# HBV and HCV prevalence and treatment eligibility assessment at a primary health facility in Ghana

**DOI:** 10.1101/2024.05.01.24306678

**Authors:** Adanusa Madison, George Adjei, Seth Agyeman, Cecil Banson, Benjamin Nyane, Rudolf Aaron Arthur, Samuel Amoah, Faustina Halm-Lai, Dennis Boadi, Oheneba Charles Kofi Hagan

## Abstract

**Background:** Viral hepatitis continues to be of global public health importance, especially hepatitis B and C. Hepatitis B and C account for most of the mortality and morbidity attributable to these viral infections. There is lack of accessibility to diagnostic tools, assessment for the eligibility for treatment and treatment in most LMICs, including Ghana. Assessment for HBV treatment is expensive and not widely available throughout Ghana. TREAT-B, a new, affordable and widely available assessment tool for HBV treatment eligibility, offers a cost-effective alternative to the international and local guidelines currently in use.

**Aim:** To determine the prevalence of HBV and HCV in a cohort of patients and to assess the HBV infected for treatment eligibility.

**Materials and methods:** A cross-sectional study of individuals diagnosed with hepatitis B infection was undertaken to assess their eligibility for treatment using local and international treatment guidelines. An additional retrospective review of records of patients screened for hepatitis B and C viruses over the period was undertaken. Questionnaires on socio-demographic were self-administered. Blood was taken for haematological analysis, liver function test, hepatitis B serological tests and DNA extraction for hepatitis B genotyping and drug sensitivity determination.

**Results:** From the retrospective review, 3358 individuals had complete data for further analysis. Prevalence for HBV and HCV was estimated at 4.3% and 1.9% respectively. Males were 3 times more likely to be HBV positive compared to females (AOR=2.8, 95% CI: 1.8-4.2, *p*-value < 0.001). Those who were born after the introduction of after the UCVHB had higher odds of having HBV compared to those who were born before (AOR=3.6, 95% CI: 2.5-5.3, *p*-value < 0.001). Assessment of 74 HBV positive patients found, 20%, 9.5%, 4.7% and 4.7% were eligible for treatment according to the TREAT-B, the WHO simplified, Ghanaian and AASLD guidelines respectively.

**Conclusion:** The burden of HBV and HCV infections remain high in Ghana and policies must be implemented to treat individuals eligible.

## Introduction

Viral hepatitis infections continue to be of public health importance globally, with approximately 2.8 million of the global population have had prior exposure to the viruses and over 1.4 million deaths recorded annually as a result (Jefferies et al., 2018). These infections are mostly caused by a group of diverse hepatotropic viruses, including Hepatitis A (HAV), B (HBV), C (HCV), D (HDV) and E (HEV) (Jefferies et al., 2018; Ustianowski & Devine, 2020). Among these viruses, HBV and HCV account for 90 - 95% of the mortalities resulting mainly from liver complications, including cirrhosis, liver failure and hepatocellular carcinoma (HCC) (Palayew et al., 2020). In 2019 alone, there were an estimated 298 million individuals globally living with chronic HBV (CHB) infection and 296 million deaths resulting from CHB complications with 1.5 million new HBV infections (World Health Organization, 2021). Similarly, approximately 1.5 million new infections were reported for HCV globally as at 2019, 58 million individuals were living with chronic HCV infection with 290 000 deaths reported in the same year (World Health Organization, 2021).

In 2015, the World Health Organisation devised an ambitious strategy of eliminating viral hepatitis, especially HBV and HCV, as a public health problem by the year 2030 (World Health Organization, 2016). Included in the 2016 WHO strategy was the reduction of mortality from HBV and HCV by 65% by 2030 from the 2015 baseline through numerous interventions including the enhancement of diagnosis outreach and the treatment of eligible individuals (World Health Organization, 2016b). However, HBV and HCV infections remain underdiagnosed globally, especially in Africa and Southeast Asia regions, where a greater burden of the infection is carried (Ginzberg et al., 2018; Hsu et al., 2023). Further, access to treatment for even the few who are diagnosed remains poor in these countries, stemming from inaccessibility to diagnostic tools for treatment eligibility assessment (Martyn et al., 2023). This phenomenon threatens to jeopardise the WHO strategy of reducing mortality from HBV and HCV by 65% globally, especially in these countries (Hsu et al., 2023).

Most countries in the AFRO region of Africa have adopted the WHO guideline for the eligibility assessment for treatment of HBV infection although the guideline was developed using data from non-African population (Johannessen et al., 2022). Unsurprisingly, this treatment algorithm has performed poorly when used to assess African patients for HBV treatment eligibility (Dusheiko & Lemoine, 2019; Johannessen et al., 2022). Additionally, the WHO guideline entails the patient to undertake ancillary laboratory investigations (full blood count, nucleic acid quantification, virus genotyping, serological profiling, liver function tests) and radiological tests (ultrasonography and liver elastography) which are inaccessible to the majority of the African populace burdened by CHB (Johannessen et al., 2019; Luong Nguyen et al., 2024). Recently, TREAT-B guideline which uses the HBV e antigen (HBeAg) status and serum ALT concentration to determine treatment eligibility for HBV treatment has been advocated (Shimakawa et al., 2018). Initially developed using individuals from The Gambia PROLIFICA project, the guideline has been evaluated in diverse populations including Europeans, Asians, Africans and Australians and has been found to have higher sensitivity and specificity compared with WHO guideline (Geeratragool et al., 2023; Howell et al., 2020; Shimakawa et al., 2019; Shimakawa & Lemoine, 2020; Vinikoor et al., 2020; Vu Hai et al., 2021). However, with the reported low specificity of the proposed guideline, the propensity for “overtreatment” could be potentially high (Luong Nguyen et al., 2024). Inasmuch, the increased accessibility using TREAT-B guideline could potentially improve the current woeful treatment rate of 1 in 1000 eligible patients in Africa (Sonderup & Spearman, 2022; Spearman et al., 2023; World Health Organization, 2021). Additionally, some studies have reported that some patients deemed ineligible for CHB treatment based on conventional guidelines may eventually developed liver complications (Han & Tran, 2015).

The Ghanaian treatment guideline developed in 2014 stipulates treatment for all patients with HBV-related cirrhosis and fibrosis (APRI > 2) with detectable viraemia and chronic active hepatitis, that is persistently elevated ALT (> 2x ULN) (ULN not defined) and viraemia of ≥ 20,000 IU/ irrespective of HBeAg status. However, A recent study at a primary health centre in the Eastern Ghana reported a significant number of patients were unable perform viral load, an essential component in the Ghanaian guideline (Duah & Nartey, 2023). Would the adoption of a simple, yet sensitive guideline like TREAT-B in Ghana, especially at the primary health care level result in reducing the burden of CHB complications?

We previously undertook a retrospective review of medical records of patient screened for HBV and HCV and reported a higher burden of HBV among individuals born before the introduction of UVHB in 2002 (Adanusa et al., 2023). The current study was undertaken to determine the burden of HBV and HCV in a cohort of students screened for the infections in 2022 and to assess their eligibility for HBV treatment based on various international and local guidelines.

## Materials and methods

### Study design

The study was a cross-sectional study with an additional retrospective review of patient medical electronic data. The study was undertaken at the University Cape Coast Hospital, a 100-bed capacity primary health facility located in the Cape Coast Metropolitan area in Ghana. The study was undertaken from February to June 2022. Patients included in the study were patients who had been screened for HBV and HCV at the hospital over the period and had tested positive to HBV. All patients who tested positive for HBsAg test and were referred for counselling and review by a physician were eligible to be recruited into the study. Written informed consent was initially sought and once granted, the individuals were subsequently enrolled into the study. The study protocol was reviewed by the University of Cape Coast Institutional Review Board and ethical clearance granted (UCCIRB/EXT/2021/04) before the study commenced.

### Procedure

#### Questionnaire administration

Questionnaires covering the participant socio-demographic status including age, sex, region of birth and residence, and parental occupation and educational status were self-administered to the participants once informed consent had been sought and granted.

#### Laboratory tests

A total of 6 ml of blood was then taken from the participant through a venepuncture, 2 ml into an EDTA sample for the haematological analysis. The remaining 4 ml was put into a serum separator tubes (SST) for the other downstream procedures including HBV profile test, liver function test, HBV viral load estimation and HBV DNA extraction. The sample in SST were allowed to stand for about 15 minutes at room temperature to clot. The clotted sample was further centrifuged at 3000g for 3 minutes and the serum aliquoted into Eppendorf tubes, stored at -20°C until further use.

### Haematological, serum chemistry and hepatitis B serological analyses

Haematological analysis was performed immediately after the venepuncture to determine the haematological indices of the participants including haemoglobin concentration (HB), platelet count, mean corpuscular haemoglobin (MCH) and mean corpuscular volume (MCV). The haematological analyses were performed on Yumizen H500 OT Haematology Analyzer (Horiba Medical, Japan)

To determine the HBV serological status of the participants, the Wondfo One Step HBV Combo Test Kit (Guangzhou Wondfo Biotech Co., Ltd, China) was utilised. The tests were performed according to the manufacturer’s protocol. Briefly, a drop each of the patient serum was put into the test kit wells designated for the specific serological marker. The test kit buffer was added to each well and the result read in 15 minutes as either positive for the marker or negative. The HBV serology tested included the HBV surface antigen (HBsAg), surface antibody (HBsAb), e protein antigen (HBeAg), e protein antibody (HBeAb) and core protein antibody (HBcAb).

The serum aspartate transaminase (AST) and alanine transaminase (ALT) concentrations were determined using the Mindray BS-120 automated Chemistry Analyzer (Shenzen Mindray Biomedical Electronic Co., Ltd, China) from the stored serum samples. Results were recorded in IU/L.

### APRI and FIB-4 estimations

APRI was estimated using the formula: [(AST/upper limit of the normal AST range) X 100]/Platelet Count (Lin et al., 2011). The upper limit of normal (ULN) used in the calculation was 40 IU/L. FIB-4 was estimated using the formula: Age ([yr] x AST [IU/L]) / ((PLT [10^9^/L]) x (ALT [IU/L])(1/2)) (Sterling et al., 2006). All calculations were performed in MS Excel (Microsoft Inc., USA).

### HBV viral load measurements

The HBV viral load measurement was undertaken using the Roche Cobas 6800 (Roche Diagnostics International, Switzerland). The tests were undertaken according to the manufacturer’s instructions. Thawed patient serum samples were loaded into the sample-supply module of the instrument. All assays were performed according to the manufacturer’s instructions. We applied both the 500µL protocol when analysing the samples.

### Hepatitis B molecular analyses

To perform the molecular analyses, DNA was initially extracted from patient serum sample using the Zymo Quick DNA extraction kits (Zymo Research, USA) according to the manufacturer’s protocol for extracting DNA from serum. The quality and quantity of DNA was determined using the Nanodrop ND-1000 UV-Vis Spectrophotometer (Thermo Fisher Scientific, USA). PCR was performed using the forward primer GATGTGTCTGCGGCGTTTTA (378-397) and the reverse primer CTGAGGCCCACTCCCATAGG (639-658) previously reported by (Astbury et al., 2020). These primers amplified a 281-nucleotide long sequence encompassing the overlapping S and P portions of the HBV genome. The reaction was performed using the OneTaq 2X master mix kits (New England Biolabs, USA). The reaction was performed in total volume of 25 uL made up of 12.5 μL of OneTaq 2X master mix, 0.5 μL (10 μM) each of the forward and reverse primers, 2 μL of DNA and 9.5 μL of nuclease free water. The cycling conditions were as follows: initial denaturation at 94°C for 180 seconds; 30 cycles of denaturation at 94°C for 30 seconds, annealing at 60°C for 40 seconds and extension of 72°C for 50 seconds; followed by final extension at 72°C for 10 minutes.

The amplicons were subsequently sent to Inqaba Biotech West Africa for the Sanger sequencing.

### Genotyping and phylogenetic analysis

The NCBI database was queried using BLASTN (https://blast.ncbi.nlm.nih.gov/Blast.cgi) to determine the genotypes of the sequences based on similarity scores. Additionally, sequences were submitted to the GENAFOR (https://hbv.geno2pheno.org) for HBV genotyping, sub genotyping and drug sensitivity prediction.

To perform the phylogenetic analysis, sequences from different HBV genotypes were downloaded from the NCBI database and HBVdb (https://hbvdb.lyon.inserm.fr/HBVdb/HBVdbIndex). The phylogenetic analysis was performed in CLC Workbench (Qiagen, Denmark). Sequences were initially aligned using default settings. Subsequently, Maximum Likelihood phylogenetic algorithm was utilised to perform the phylogenetic analysis on the previously aligned sequences utilising default settings based on 1000 bootstraps pseudo-replicates.

### Statistical analyses

Sero-prevalence was calculated as the percentage of participants with any type of infection for each sub-category. Pearson’s chi-squared was utilised to determine the association between participant characteristics and infection status. Multivariable logistics modelling was used to determine the risk factors associated with infection status. The results were expressed as Odds ratios with 95% confidence interval. A *p*-value of less than 0.05 was considered significant for all the statistical analyses. All statistical analyses were undertaken using the STATA 14 statistical analytical tool (Stata Corporation, USA).

## Results

### Sociodemographic characteristics and predictors of HBV and HCV infections

Three thousand four hundred and forty (3440) individuals were screened for HBV and HCV during the study period. Out of this number, 3358 had complete data for further analysis. The participants had a median age of 20 (IQR = 19, 22). Of these, 1715 (51.1%) identified as males with median age of 21 (IQR = 19.0, 22.0) and 1643 (48.9%) identified as females with a median age of 19 (IQR = 18, 21). Majority of the individuals, 57.6%, were born after 2002, the year Ghana introduced the universal child vaccination against HBV (UCVHB) into the expanded programme of immunisation (EPI).

Of the 3359 individuals with complete data, 145 of them tested positive for HBV, a crude prevalence of 4.3%. The prevalence in the male population was 6.6% (113 of the 145) and in the female population was 2.0% (32 of the 145). The prevalence in the individuals born after the introduction of the UCVHB was 1.9% (36 of the 145) and 7.7% (109 of 145) in those born before. Of the number screened with complete data, 62 tested positive for HCV, a proportion of 1.9%. The prevalence in the males was 1.8% (31 positive) and 1.9% (31 positive) in the females. In the age category 20 years or below group, the prevalence was 1.76% (34 positive) and 2.0% (28 positive) in those above 20 years of age.

To determine the risk factors associated with infection with HBV and HCV and socio-demographic characteristics, we performed a multi-variate logistics regression analysis to determine the crude and adjusted Odds ratio. The result is presented in table 1. In the multivariable logistic regression, males were 3 times more likely to be HBV positive compared to females (AOR=2.8, 95% CI: 1.8-4.2, *p*-value < 0.001). Those who were born after the introduction of after the UCVHB had higher odds of having HBV compared to those who were born before (AOR=3.6, 95% CI: 2.5-5.3, *p*-value < 0.001). In the multivariable logistic regression, males were 7% less likely to be HCV positive compared to females (AOR=0.9, 95% CI: 0.6-1.6, *p*-value = 0.787). Those who were introduced after UVC had higher likelihood of having HCV compared to those who were introduced before UVC (AOR=1.14, 95% CI: 0.68-1.91, *p*-value = 0.624) (Table 2). However, none of the variables was associated with HCV status in the multivariable regression.

**Table 1.** Table presents the factors that influence HBV status. In the multivariable logistic regression. * also represents participants 20 years or younger.

| Variable | Sub-category | OR<br>(95% CI) | $p$-value | AOR<br>(95% CI) | $p$-value |
| --- | --- | --- | --- | --- | --- |
| **Sex** | Female | 1 (ref) |  | 1 (ref) |  |
|  | Male | 3.6<br>(2.4-5.3) | <0.001 | 2.8<br>(1.8-4.2) | < 0.001 |
| **UVC\*** | Before UVC | 1 (ref) |  | 1 (ref) |  |
|  | After UVC | 4.4<br>(3.0 -6.4) | <0.001 | 3.6<br>(2.5-5.3) | < 0.001 |

**Table 2.** Table depicts the factors that influence HCV status of participants.

| **Variable** | **Sub-category** | **OR**<br>**(95% CI)** | **p-value** | **AOR**<br>**(95% CI)** | **p-value** |
| --- | --- | --- | --- | --- | --- |
| **Sex** | Female | 1 (ref) |  | 1 (ref) |  |
|  | Male | 0.96<br>(0.6-1.6) | 0.86 | 0.93<br>(0.6-1.6) | 0.787 |
| **Age** | **<= 20** | 1 (ref) |  | 1 (ref) |  |
|  | > 20 | 1.1<br>(0.7-1.9) | 0.658 | 1.1<br>(0.7-1.9) | 0.624 |

### Eligibility assessment

Of the 140 individuals who tested positive for HBV during the screening, 77 consented to be part of the eligibility assessment study. Two (2) tested negative for HBsAg and HBcAb when retested and were, therefore, excluded from the study. One (1) individual did not have LFT results and was also excluded from further assessment. The remaining 74 individuals had a median (IQR) age of 23.0 (20.0, 26.8) years. Out of the 74, 78.4% (58) were males with a median age of 22.0 (IQR = 20.0, 26.0) years and 21.6% (16) identified as females with a median age of 23 (IQR = 20.3, 25.8) years.

The haemoglobin concentration (HB), platelet count, mean corpuscular haemoglobin (MCH), mean corpuscular volume (MCV), ALT, AST, APRI, FIB-4 and HBV viral load results are presented in Table 3 as median (IQR). Viral load results are for 64 individuals, for whom the result was available at a minimum cut of 20 IU/mL. Three (3) individuals tested positive for HBeAg.

**Table 3.** Table showing the medians of laboratory test results for the HBV positive participants and the IQRs.

| **Laboratory test** | **Median (IQR)** |
| --- | --- |
| HB (g/dL) (N = 74) | 14.0 (13.0, 14.8) |
| Platelet count (N = 74) | 207.5 (168.8, 239.3) |
| Haematocrit (%) (N = 74) | 42.6 (39.6, 45.6) |
| ALT (IU/L) (N = 74) | 25.0 (19.0, 36.0) |
| AST (IU/L) (N = 74) | 29.5 (23.0, 34.5) |
| MCV (N = 74) | 86.9 (82.7, 90.5) |
| MCH (N = 74) | 28.3 (26.9, 30.0) |
| APRI (N = 74) | 0.4 (0.3, 0.5) |
| FIB-4 (N = 74) | 0.7 (0.6, 0.9) |
| HBV Viral load (IU/L) (N = 64) | 872.0 (45.3, 5615.0) |

Assessing the patients for treatment eligibility using the different guidelines, 20% (15 out of 74) were eligible for treatment according to the TREAT-B guideline as shown in Figure 1. The WHO simplified guideline had 9.5% (7 out of 74) eligible for treatment, with 4.7% each being eligible for treatment according to the Ghanaian and AASLD guidelines. None of the participants assessed was eligible for treatment based on the WHO guideline. Using the APRI cut-off of ≥ 2 as a proxy for the cirrhosis, none of the participants was diagnosed with cirrhosis.

**Figure 1.**
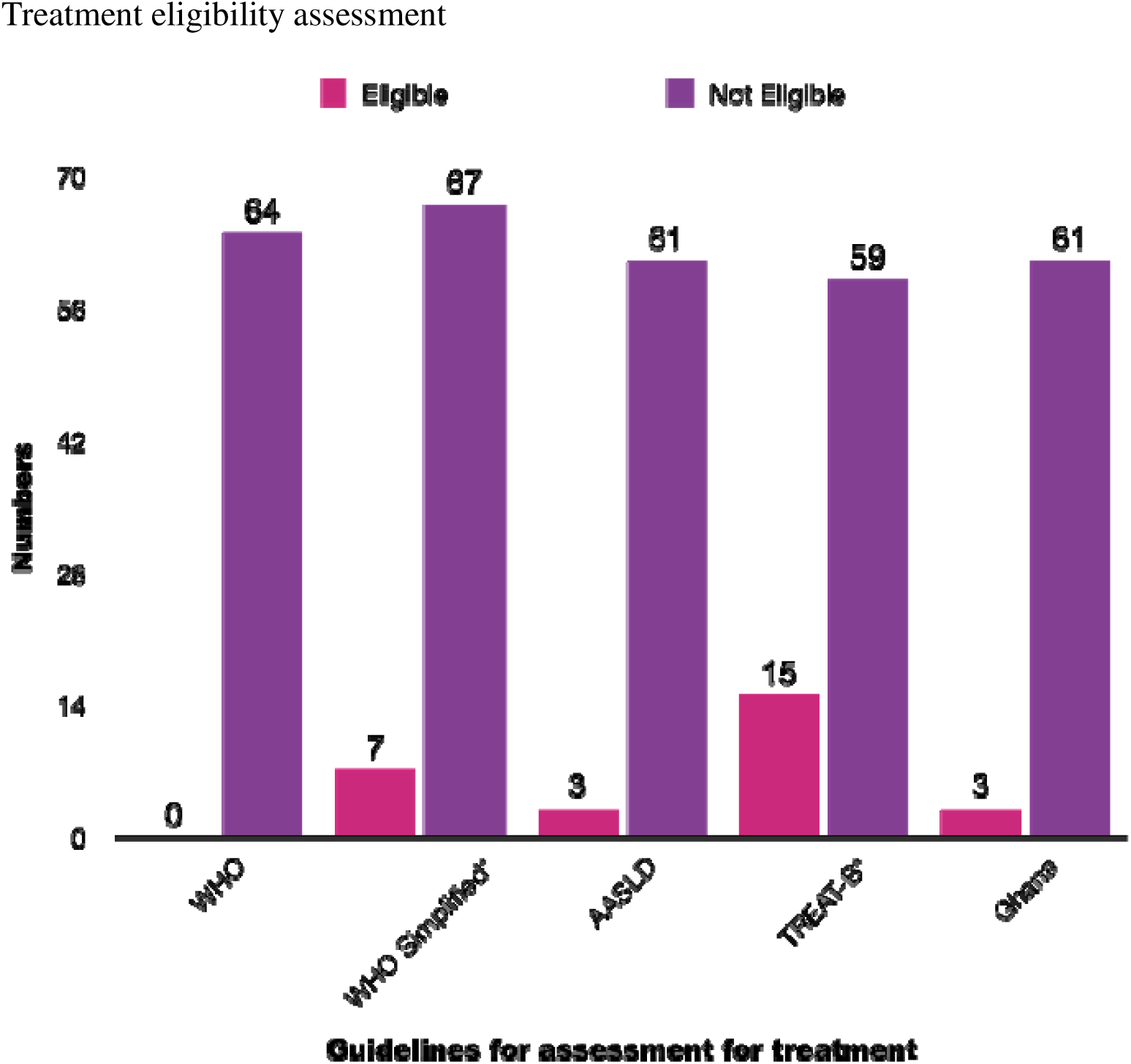
Bar chart showing the number of individuals who were eligible for HBV treatment according to different treatment guidelines and those who were ineligible. * 74 individuals assessed

### Genotyping and phylogenetic analysis

We successfully amplified and sequenced 4 of the samples. Genotyping with the GENAFOR algorithm identified 3 of the samples as belonging to genotype E and the remaining as genotype A sub genotype A1 shown in Table 4. All the 4 samples showed sensitivity all the commonly used drugs for HBV treatment. Table 4 shows the genotype and drug sensitivity profile of the sequenced samples.

**Table 4.** shows the genotype and drug sensitivity genotyping for successful PCR-amplified HBV.

|  | Genotype | Sub-genotype | Lamivudine | Adefovir | Entecavir | Tenofovir<br>DF | Viral Load<br>(IU/mL) | TREAT-B<br>eligibility |
| --- | --- | --- | --- | --- | --- | --- | --- | --- |
| HB18 | E | N/A | Sensitive | Sensitive | Sensitive | Sensitive | 20 | No |
| HB32 | E | N/A | Sensitive | Sensitive | Sensitive | Sensitive | 60096 | No |
| HB43 | A | A1 | Sensitive | Sensitive | Sensitive | Sensitive | 6019 | No |
| HB53 | E | N/A | Sensitive | Sensitive | Sensitive | Sensitive | 20 | No |

The phylogenetic analysis was performed for the sequences against known specific genotype sequences shown in Figure 2.

**Figure 2.**
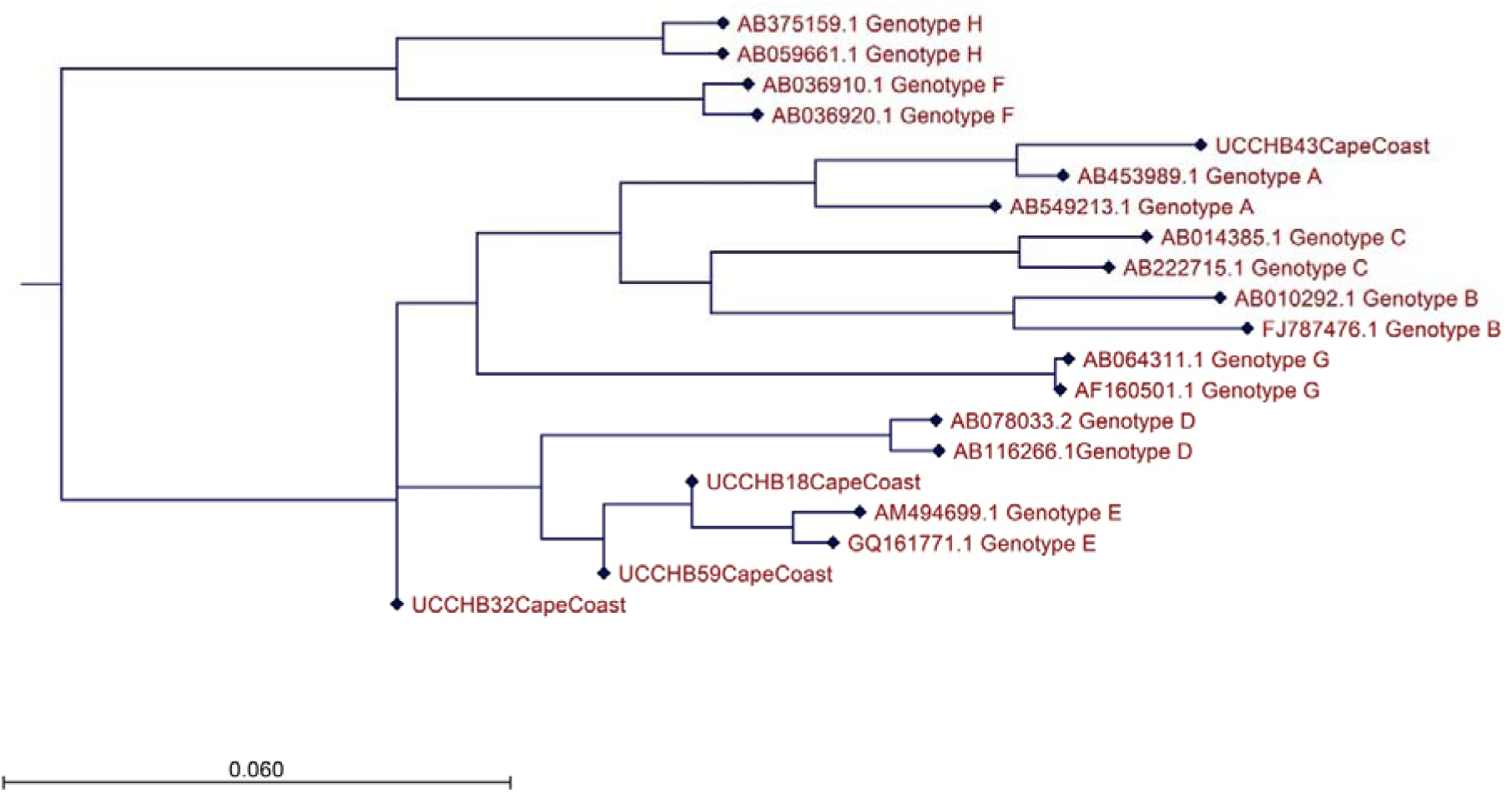
Bootstrap consensus Maximum Likelihood phylogenetic tree of the samples and known HBV genotypes accessed from the HBVdb database.

## Discussion

The WHO strategy to eliminate viral hepatitis as a public health problem by 2030 hinges on strategies for reducing mortality from HBV and HCV by 65% from the 2015 baseline. To achieve this goal, increased surveillance and diagnosis followed by improved linkage to health care centres for treatment and improved rollout of treatment to the eligible. The WHO goal of identifying 90% of the CHB population by 2030 might be unattainable at the current rate of 10% globally and < 1% in Africa (Sonderup et al., 2020; Spearman et al., 2023). The situation in Ghana is similar, according to the Polaris Observatory Network, as at 2022 Ghana had a CHB population of 2,769,000, of which only 0.7% had been diagnosed (Polaris Observatory Collaborators et al., 2023). Most HBV patients in Ghana and most African countries would remain undiagnosed because CHB is mostly asymptomatic and patients would usually present late to the hospital CHB complications arise (Duah & Nartey, 2023; Spearman et al., 2023). In Ghana, HBV screening is usually undertaken at health facilities, mostly based on risk assessment and supplemented by community-based testing undertaken by non-governmental organisations (Nartey et al., 2022). However, the lack of a concise national policy on viral HBV surveillance and channels of reporting threatens the goal of diagnosing at least 90% of people living with CHB by 2030 (Coalition for Global Hepatitis Elimination, 2022).

The crude prevalence rate for HBV infection in our study is lower than the reported aggregated seroprevalence of 12.3% (Ofori-Asenso & Agyeman, 2016). The declining prevalence in Ghana has been attributed to HBV infection prevention policies in Ghana, especially the introduction of UCVHB into the EPI and mother-to-child prevention strategies (Ghana Ministry of Health, 2016). Nonetheless, the prevalence of HBV before the introduction of UCVHB remains high, meaning the burden from complications of CHB would be expected to increase in this cohort in the foreseeable future. It has, therefore, become imperative that health authorities increase the effort to target individuals in the “before UCVHB” cohort for diagnosis, eligibility for treatment assessment and treatment. The current study sought to assess individuals positive for HBsAg for eligibility for treatment based on the current Ghanaian, WHO, TREAT-B and AASLD guidelines. For individuals whom eligibility could be performed in our study, a higher proportion were eligible for treatment based on the TREAT-B guideline compared with the other guidelines. The TREAT-B eligible proportion of 20% is comparable to studies on TREAT-B eligibility reported by a community-based study in Australia and a hospital-based study in Ethiopia (Howell et al., 2020; Johannessen et al., 2019). However, the proportion eligible for treatment based on TREAT-B was lower compared to studies undertaken in Zambia, Vietnam, Uganda, Burkina Faso, The Gambia and Australia, the proportion of eligible patients for treatment were higher than the current study a higher proportion (Howell et al., 2020; Kafeero et al., 2022; Shimakawa et al., 2019; Vinikoor et al., 2020; Vu Hai et al., 2021). The proportion of TREAT-B eligible individuals in the current study was lower likely because the population was younger and mostly from the community compared with the other studies. The need for treatment would be expected to be higher in hospital-based studies because patients are likely to be CHB symptomatic and would be tested based on risk assessment compared with participants detected in community-based surveillance and are likely to be CHB asymptomatic. However, the high proportion reported by the authors (Vinikoor et al., 2020), although community-based, can be attributed to an older population compared to a much younger population. Additionally, factors such as HBV genotype influence disease progression and may impact on eligibility for treatment (Sonderup et al., 2020). For example, Genotype E predominantly found in West Africa is associated with high HBeAg carriage coupled with high viral load and Genotype A1 also found in some African regions, especially Eastern Africa, is associated with early loss of HBeAg and a faster progression to CHB complications such as HCC (Sonderup et al., 2020; Thurnheer et al., 2017). Although we could not genotype all the HBV in our study, 3 out the 4 genotyped were genotype E which predominates in Ghana (Anabire et al., 2023; Candotti et al., 2006; Dongdem et al., 2016). This agrees with our study with the low HBeAg positivity proportion in our study, which may need further study to elucidate the effect of the genotypes on disease progress in Ghana.

A recent study on the economic benefit of treating HBV has shown that, the adoption of TREAT-B guideline for treatment in LMICs especially countries in Africa would be cost-effective compared to alternative treatment options (Luong Nguyen et al., 2024). Additionally, treatment can be undertaken even at primary health care facilities as the HBeAg and ALT can be performed easily and treatment can be initiated immediately overcoming the multiple follow-ups prior to treatment for the other guidelines. Nonetheless, TREAT-B is currently not an accepted guideline because the scant available evidence in addition to low specificity which can result in over-treatment and current point of care tools for HBeAg determination are currently unreliable (Stockdale et al., 2021). An expert opinion based on a study in Ethiopia has recommended lowering the APRI threshold from ≥ 2 to ≥ 0.65 (Spearman et al., 2023). Interestingly, none of the participant in our study participants were eligible for treatment based on the WHO threshold, however, when the cut-off was lowered to a threshold of ≥ 0.65, 7 patients would have been diagnosed with cirrhosis. Thus, the combination of APRI at a cut-off of ≥ 0.65 for cirrhosis assessment and the TREAT-B guideline would increase proportions eligible for treatment, albeit at the expense of lowering the specificity.

According to the Polaris Observatory Network, there are currently an estimated 440,000 people living with chronic HCV infection in Ghana, 9% of whom have been diagnosed with less than 1% treated annually. Previous systematic review reported HCV prevalence of 3% in Ghana with a recent nationwide auditing of hospital records estimating the prevalence at 4.6% (Agyeman et al., 2016; Nartey et al., 2023). The proportion from our study with HCV seropositive of 1.85% was less than these estimates but closer to the prevalence of 2.7% reported by a study undertaken previously in Cape Coast, the locale for our study (Tetteh et al., 2020). Because 30% of HCV serologically positive patients do not need treatment, treatment eligibility is based on detection of HCV ribonucleic acid (RNA) in patient serum (Patel et al., 2021). Because the HCV RNA determination was not undertaken in this study, we were unable to determine the proportion of the HCV positive individuals who would have needed treatment. The cost of viral load estimation in Ghana is high, with ancillary tests including HCV genotyping and liver fibrosis status (using FIB-4, APRI, liver elastography and liver biopsy) adding up additional cost to the patients (Nartey et al., 2023). The introduction of the curative DAAs for HCV treatment makes it imperative that population screening ought to be scaled up especially among high-risk groups, including incarcerated individuals, people who inject drugs and children born to mothers with HCV. The recent launch of Screening and Treatment Opportunity Project for Hepatitis C (STOP Hep C) by Ghana Ministry of Health in collaboration with government of Egypt to provide treatment with DAA for HCV patients nationwide is auspicious (Ghana Health Service, 2023). Nonetheless, the cost of liver fibrosis, viral RNA and HCV genotyping prior to treatment initiation could remain an obstacle to treatment.

Accessibility to treatment for viral hepatitis especially HBV and HCV remain a challenge in LMICs including Ghana. Underdiagnosis of infected individuals coupled with lack of eligibility assessment for treatment due to high attrition rate of those diagnosed and expensive drugs for treatment. At the primary health care facilities, these challenges become exaggerated, however, integration of diagnosis, assessment for treatment and treatment at these facilities has been proposed to increase accessibility. TREAT-B guidelines for HBV and the STOP Hep C for HCV when coordinated with screening for viral hepatitis at primary health care facilities would improve the chances of Ghana achieving the WHO 2030 targets.

## Limitations

The sample size for the eligibility for HBV treatment was small compared to similar studies undertaken elsewhere. Because the more sensitive liver biopsy and elastography were not performed, we could not determine the performance of TREAT-B guidelines in Ghanaian compared to the gold standards employed in studies elsewhere. Therefore, the utility of TREAT-B guideline for treatment assessment in Ghanaian population cannot be ascertained. Similarly, we could not determine eligibility for HCV treatment due to inadequate funding.

## Data availability

The data from which the manuscript was derived has been deposited in the Mendeley Data (Hagan, Oheneba C K (2024), “Cape Coast Hepatitis B and C Screening and treatment eligibility assessment dataset”, Mendeley Data, V1, doi: 10.17632/4d6xynfdsy.1) Sequences from the Sanger sequencing has been deposited in the Genbank.

